# Genome-Wide DNA Methylation Profiling Reveals Enrichment of Developmental Vascular and Muscle Programs in Peripheral Artery Disease: A Pilot Epigenome-Wide Association Study

**DOI:** 10.64898/2026.08.03.26359321

**Authors:** Aditya Safaya, Kristen Spreha, Christina Jones, Victor Ruiz-Velasco, Piotr K. Janicki

## Abstract

**Background:** Peripheral artery disease (PAD) is a common atherosclerotic condition with incompletely understood molecular drivers. DNA methylation is a key epigenetic regulator influenced by smoking, aging, and metabolic risk factors, yet genome-wide methylation patterns specifically associated with PAD remain understudied.

**Objective:** To characterize genome-wide DNA methylation patterns associated with peripheral artery disease (PAD) using the Infinium Methylation EPIC v2.0 platform.

**Design:** Single-center pilot epigenome-wide association study using pooled whole blood genomic DNA and pool-level bioinformatic analysis. Setting: Academic medical center. Patients: Ten adults patients diagnosed with PAD and 22 healthy controls.

**Interventions:** None.

**Methods:** In this prospective single-center pilot study, we performed genome-wide DNA methylation profiling on blood DNA from 10 clinically diagnosed patients with PAD (mean age 72.7 ± 11.2 years; 50% female) and 22 age- and sex ratio-matched controls (mean age 74.6 ± 7.8) using the Illumina Infinium MethylationEPIC v2.0 BeadChip (>935,000 CpG sites). Data was processed with the SeSAMe pipeline. Differentially methylated loci (DMLs) and regions (DMRs) were identified at FDR ≤ 0.01 and |Δβ| ≥ 0.10. Enrichment analysis of associated genes was performed using clusterProfiler (Gene Ontology, GO, and Kyoto Encyclopedia of Genes and Genomes, KEGG, computational methods).

**Results:** Following quality control, 933,942 probes were analyzed. We observed that a majority of significant DMLs exhibited hypomethylation in PAD. Intersection of DML- and DMR-supported genes yielded 16,854 high-confidence genes. GO analysis revealed a strong enrichment for muscle system processes, regulation of membrane potential, embryonic organ development, DNA-binding transcription activator activity, actin binding, and metal ion transmembrane transporter activity. Cellular component terms highlighted cell leading edge, focal adhesion, and cell cortex. KEGG pathways were dominated by calcium signaling, cadherin signaling, MAPK signaling, and cytoskeleton in muscle cells.

**Conclusions:** Blood DNA methylation patterns in a cohort of PAD patients are enriched in developmental vascular, muscle, and cell-motility programs that are characteristically reactivated in adult vascular pathology (endothelial dysfunction, vascular smooth muscle cell (VSMC) phenotypic switching, pathological angiogenesis). These findings generate a hypothesis for biomarker development and epigenetic therapeutic targeting in PAD. Larger longitudinal and multi-ethnic validation studies are warranted.

## Introduction

Peripheral artery disease (PAD) is characterized by atherosclerotic narrowing or occlusion of arteries supplying the lower extremities, resulting in reduced tissue perfusion, claudication, impaired wound healing, and elevated risk of major amputation and cardiovascular events. PAD shares classical risk factors with other atherosclerotic diseases—including smoking, diabetes mellitus, hypertension, dyslipidemia, and advanced age—yet its molecular pathogenesis remains incompletely defined.

Epigenetic mechanisms, particularly DNA methylation (the covalent addition of methyl groups to cytosine residues, predominantly at CpG dinucleotides), provide a critical interface between environmental exposures and gene regulation. Methylation can silence or modulate gene expression without altering the underlying DNA sequence and is highly modified by inflammation, smoking, chronological aging, and metabolic stress—all risk factor for PAD. Differentially methylated loci (DMLs) and regions (DMRs) identified through epigenome-wide association studies (EWAS), therefore, can serve as potential disease biomarkers, provide mechanistic clues, and point to therapeutic targets.

Although the epigenetics of coronary artery disease and atherosclerosis have been extensively studied, dedicated genome-wide methylation analyses focused on PAD are still limited (1-5). Existing literature highlights several recurring themes: extensive hypomethylation within atherosclerotic plaques (notably at the imprinted 14q32 locus harboring a cluster of miRNAs), blood methylation signatures that largely reflect cumulative cardiovascular risk-factor exposure (especially smoking), race-associated differences in methylation landscapes, macrophage phenotypic shifts driven by promoter methylation in ischemic muscle models, and hypoxia-related gene regulation in PAD skeletal muscle. Collectively, these observations suggest that DNA methylation contributes to impaired angiogenesis, chronic inflammation, vascular remodeling, and smooth-muscle-cell phenotypic switching in PAD, yet causal and PAD-specific patterns remain poorly understood.

To address this gap, we conducted a carefully controlled pilot epigenome-wide association study comparing peripheral-blood DNA methylation profiles of clinically diagnosed patients with PAD to age- and sex-matched controls free of PAD. Using the high-density Illumina MethylationEPIC v2.0 array and rigorous statistical filtering, we sought to identify high-confidence DMLs/DMRs and to characterize the biological pathways most strongly associated with PAD status.

## Methods

### Study Design and Participants

This prospective, single-center pilot study enrolled adults (≥18 years) with a clinical diagnosis of PAD established by a board-certified vascular surgeon at Penn State Hershey Medical Center (Hershey, PA, USA) during the pre-operative evaluation for revascularization or other vascular procedures. Potential participants were first identified via electronic medical record screening and then confirmed by detailed chart review by a vascular surgeon co-author (A.S.). A total of 10 patients with PAD were enrolled. The control cohort was comprised of 22 age- and sex-ratio-matched individuals without known PAD whose de-identified genomic DNA was obtained commercially (Polbiomed LLC) with explicit consent for research use. The study was conducted in accordance with the Declaration of Helsinki; institutional review board approval and informed consent procedures were obtained as required for the clinical cohort.

### DNA Extraction and Genome-Wide Methylation Profiling

Genome-wide DNA methylation profiling was performed by TruDiagnostic Inc. (Lexington, KY, USA) using the Infinium MethylationEPIC v2.0 BeadChip array (Illumina Inc., San Diego, CA, USA). This array interrogates more than 935,000 CpG sites distributed across gene bodies, promoters, enhancers, and other regulatory elements. Genomic DNA underwent bisulfite conversion with the EZ DNA Methylation Kit (Zymo Research) according to the manufacturer’s protocol. Converted DNA was then amplified, fragmented, hybridized to BeadChips, and stained following standard Illumina workflows. Arrays were scanned on an Illumina HiScan SQ system, and raw intensity data were exported as IDAT files.

### Bioinformatic Processing and Differential Methylation Analysis

IDAT files were processed with the SeSAMe package (R/Bioconductor) (6). Quality control retained samples in which >90% of probes met a P-value threshold of 0.05. Data was normalized with the openSesame workflow under default parameters. Probes with low detection performance, those overlapping known single-nucleotide polymorphisms, sex-chromosome probes, and cross-reactive probes were excluded. Only CpG sites with <80% missing data across samples were retained for analysis. After filtering, 933,942 probes remained from the original 32 samples (all of which passed QC).

Differentially methylated loci (DMLs) were identified by two-group comparison (PAD vs. control). Statistical significance was assessed with Benjamini–Hochberg false-discovery-rate (FDR) correction. To prioritize robust signals in this pilot design, we applied stringent secondary filters: FDR ≤ 0.01 and absolute difference in methylation β-value (|Δβ|) ≥ 0.10. Differentially methylated regions (DMRs) were called with the DMR() function in SeSAMe, which groups adjacent CpGs by Euclidean distance and aggregates P-values. Two complementary gene lists were generated: (i) genes linked to individual significant DMLs and (ii) genes associated with significant DMRs. The intersection of these two lists (genes supported by both DML and DMR evidence) was used for all downstream enrichment analyses and is hereafter referred to as the high-confidence gene set.

CpG sites were annotated with the IlluminaHumanMethylationEPICv2anno. 20a1.hg38 package (gene symbols, genomic coordinates, CpG island/shore/shelf/open-sea context). Intergenic sites not mapping within 1.5 kb of a transcription start site were labeled “intergenic.” Functional enrichment analysis of the high-confidence gene set was performed with the clusterProfiler package (7), testing GO Biological Process (BP), Molecular Function (MF), and Cellular Component (CC) terms as well as KEGG pathways. Semantic similarity between cutoffs and adjusted P-value thresholds are reported with the results.

### Statistical Considerations

With an effective sample size of 10 PAD versus 22 control samples (32 observations, df = 30 before multiple-testing correction), it should be noted that this study is underpowered for genome-wide discovery and should be regarded as hypothesis-generating. All reported associations, therefore, require independent validation in larger cohorts.

## Results

### Participant Characteristics

The PAD cohort (n = 10) had a mean age of 72.7 ± 11.2 years and comprised five men and five women. The control cohort (n = 22) was closely matched ((mean age 74.6 ± 7.8) years; equal sex distribution). All participants were European ancestry.

### Widespread Differential Methylation in PAD

Of the 933,942 probes that were retained following quality control measures, 754,273 loci showed nominal differential methylation. After Euclidean-distance segmentation, 150,670 differentially methylated segments were identified, of which 92,696 remained significant after Benjamini–Hochberg correction. Application of the dual filter (FDR ≤ 0.01 and |Δβ| ≥ 0.10) yielded large gene lists: 17,306 significant genes by DML annotation and 23,309 by DMR annotation. The intersection of these two approaches produced a high-confidence set of 16,854 unique genes (Table 1). Figure 1 is a volcano plot which indicates that the majority of significant DMLs exhibited hypomethylation in PAD relative to controls.

**Table 1.**
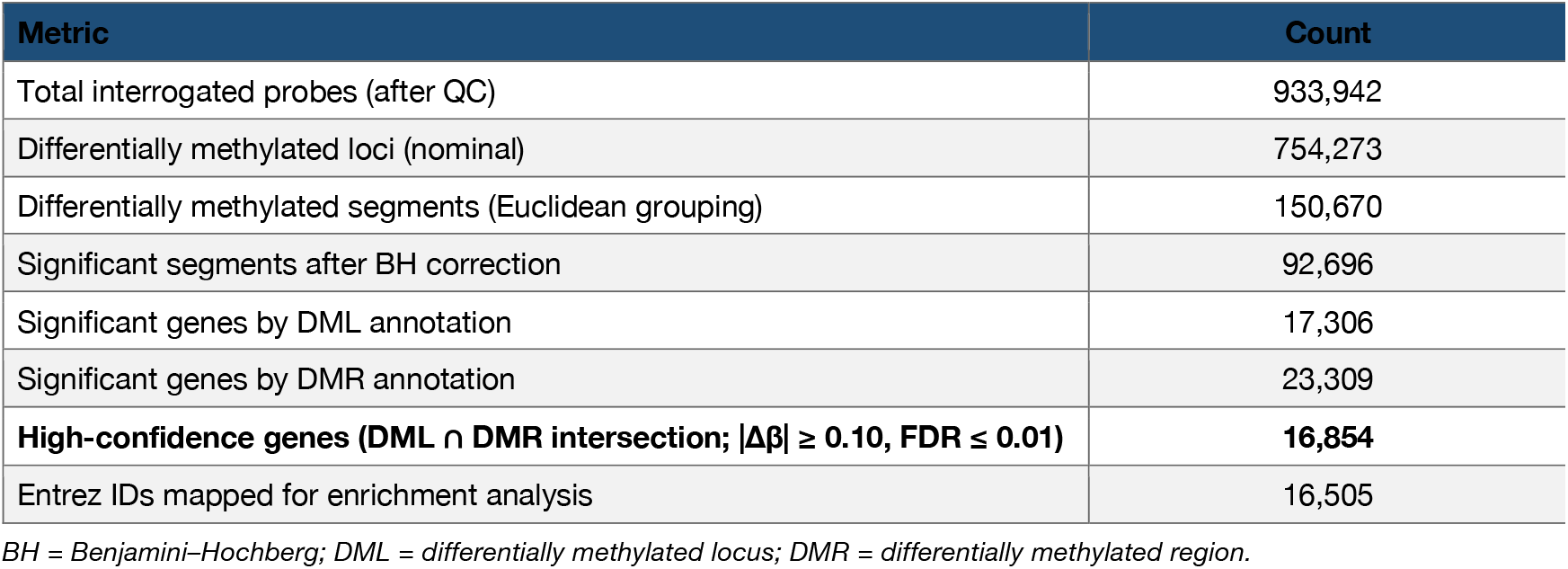
Summary of SeSAMe analysis workflow (EPIC v2.0, 32 samples)

**Figure 1.**
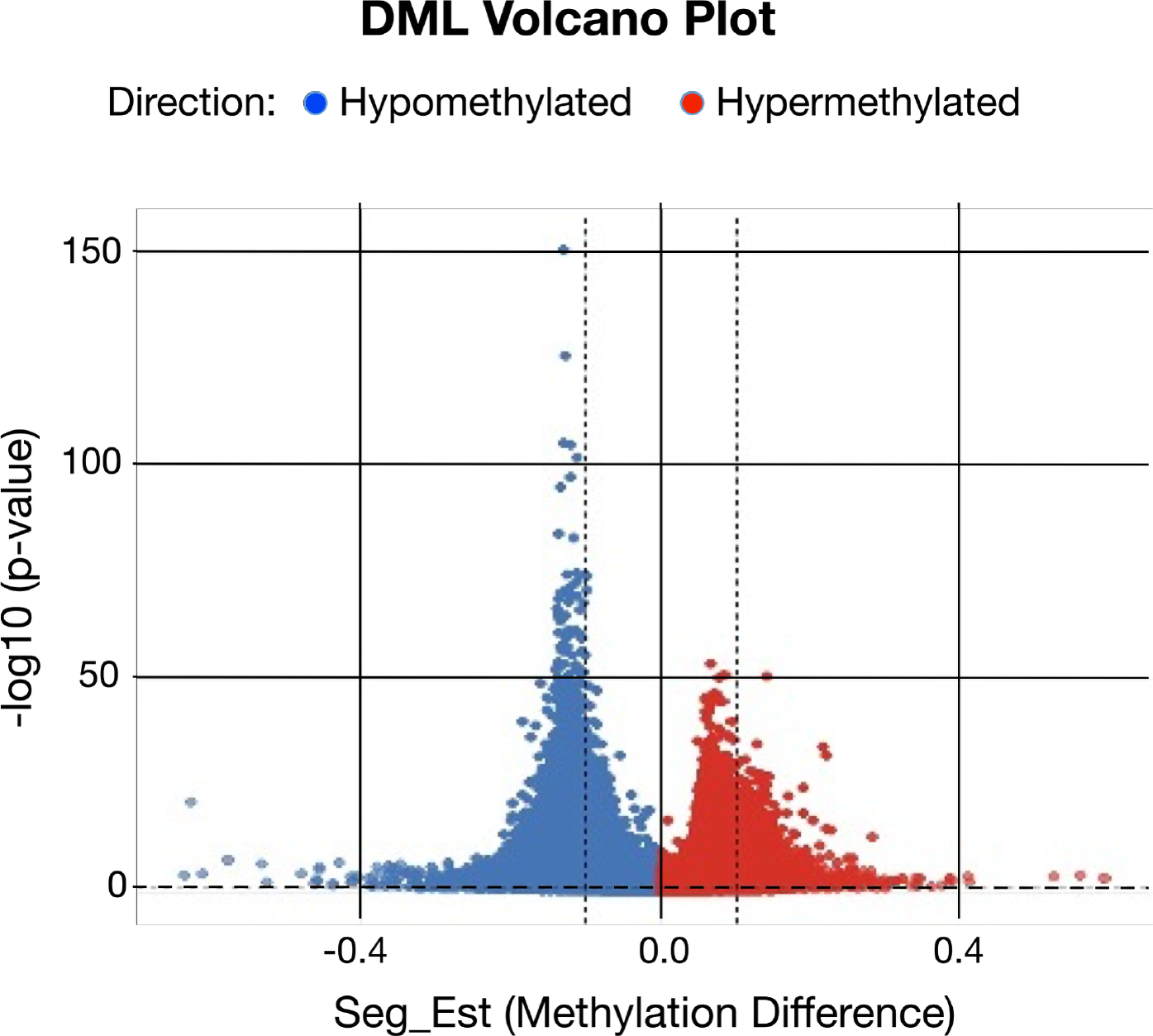
Volcano plot of differentially methylated loci (DMLs) between PAD patients (n = 10) and age-/sex-matched controls (n = 22). The x-axis shows Δβ (PAD – control); the y-axis shows – log_10_(FDR). Points are colored by direction of change (hypomethylation in blue, hypermethylation in red). Horizontal dashed line indicates FDR = 0.01; vertical dashed lines indicate |Δβ| = 0.10. The majority of significant DMLs are hypomethylated in PAD.

**Figure 2.**
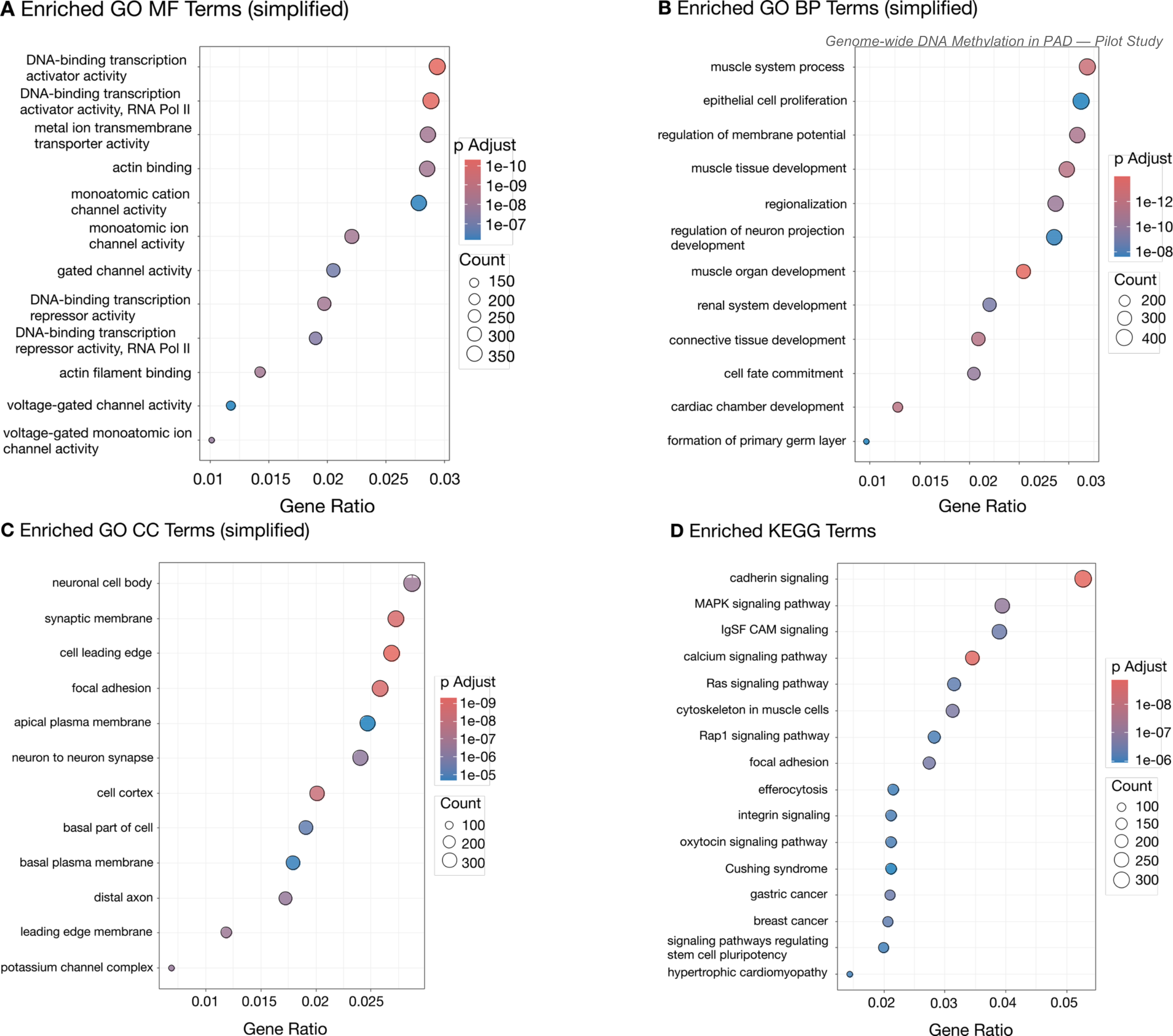
Dot plots / bar plots of the top enriched GO (BP, MF, CC) and KEGG pathways for the high-confidence gene set. Node size reflects gene count; color scale indicates adjusted P-value.

### Gene Ontology (GO) Enrichment: Reactivation of Developmental Vascular and Muscle Programs

Functional enrichment of the high-confidence gene set revealed a coherent and biologically plausible signature. Twenty-nine GO Biological Process terms survived adjusted P < 0.01 (semantic similarity cutoff 0.4; average fold enrichment ≈ 1.25). The top three terms were embryonic organ development (GO:0048568; 399 genes; Padj = 3.27 × 10^-15^), muscle system process (GO:0003012; 403 genes; Padj = 2.31 × 10^-13^), and regulation of membrane potential (GO:0042391; 391 genes; Padj = 3.06 × 10^-12^) (Table 2).

**Table 2.** Top Gene Ontology Biological Process (BP) terms.

| Rank | GO ID | Description | Count | Padj |
| --- | --- | --- | --- | --- |
| 1 | GO:0048568 | Embryonic organ development | 399 | 3.27e-15 |
| 2 | GO:0003012 | Muscle system process | 403 | 2.31e-13 |
| 3 | GO:0042391 | Regulation of membrane potential | 391 | 3.06e-12 |
| 4 | GO:0043434 | Response to peptide hormone | 368 | 2.98e-08 |

Molecular Function analysis (14 terms, semantic similarity cutoff 0.5) was dominated by DNA-binding transcription activator activity (GO:0001216; Padj = 4.63 × 10^-12^) and RNA-polymerase-II-specific transcription activator activity (GO:0001228; Padj = 5.22 × 10^-12^), together with metal-ion transmembrane transporter activity and actin binding (both Padj = 1.31 × 10^-8^) (Table 3).

**Table 3.**
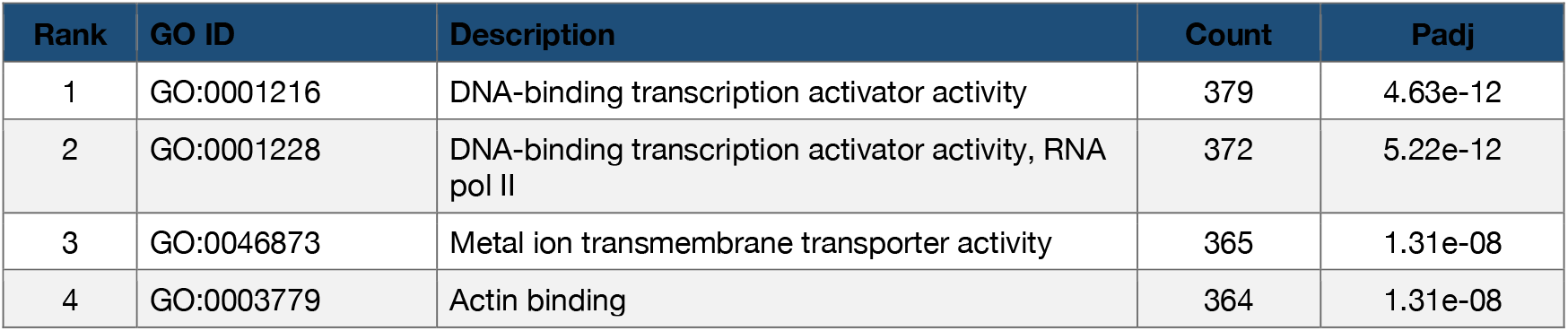
Top Gene Ontology Molecular Function (MF) terms.

| Rank | GO ID | Description | Count | Padj |
| --- | --- | --- | --- | --- |
| 1 | GO:0001216 | DNA-binding transcription activator activity | 379 | 4.63e-12 |
| 2 | GO:0001228 | DNA-binding transcription activator activity, RNA pol II | 372 | 5.22e-12 |
| 3 | GO:0046873 | Metal ion transmembrane transporter activity | 365 | 1.31e-08 |
| 4 | GO:0003779 | Actin binding | 364 | 1.31e-08 |

Cellular Component terms (13 significant) were enriched for cell leading edge, focal adhesion, cell cortex, and neuronal cell body/synaptic membrane structures (all Padj < 10^-8^) (Table 4), consistent with a cell-motility and adhesion program.

**Table 4.** Top Gene Ontology Cellular Component (CC) terms.

| Rank | GO ID | Description | Count | Padj |
| --- | --- | --- | --- | --- |
| 1 | GO:0031252 | Cell leading edge | 358 | 2.20e-10 |
| 2 | GO:0005925 | Focal adhesion | 345 | 3.50e-10 |
| 3 | GO:0005938 | Cell cortex | 270 | 5.33e-10 |
| 4 | GO:0043025 | Neuronal cell body | 384 | 4.04e-08 |

### KEGG Pathway Analysis

KEGG analysis identified 108 pathways at adjusted P < 0.01 (average fold enrichment ≈ 1.25). The leading pathways were cadherin signaling (hsa04514; 328 genes; Padj = 1.33 × 10^-9^), calcium signaling (hsa04020; 215 genes; Padj = 1.81 × 10^-9^), MAPK signaling (hsa04010; 245 genes; Padj = 8.60 × 10^-8^), and cytoskeleton in muscle cells (hsa04820; 194 genes; Padj = 1.78 × 10^-7^) (Table 5).

**Table 5.** Top KEGG pathways.

| Rank | ID | Description | Count | Padj | Category |
| --- | --- | --- | --- | --- | --- |
| 1 | hsa04514 | Cadherin signaling | 328 | 1.33e-09 | Env. Info. Processing |
| 2 | hsa04020 | Calcium signaling pathway | 215 | 1.81e-09 | Signal transduction |
| 3 | hsa04010 | MAPK signaling pathway | 245 | 8.60e-08 | Signal transduction |
| 4 | hsa04820 | Cytoskeleton in muscle cells | 194 | 1.78e-07 | Cellular Processes |

*Note: Pathway IDs updated to current KEGG nomenclature where applicable (cadherin signaling is hsa04514).*

## Discussion

In this pilot epigenome-wide association study of carefully matched PAD patients and controls, we observed extensive differential DNA methylation, with a clear predominance of hypomethylation. The high-confidence gene set supported by both DML and DMR evidence is strongly enriched for a coordinated developmental cell-adhesion, motility, and cytoskeletal program. This signature encompasses: (1) transcriptional activators that drive embryonic organogenesis; (2) components of focal adhesions and the cell leading edge that mediate migration and morphogenesis; (3) actin-binding proteins and cadherin-signaling pathways; (4) calcium signaling and membrane-potential regulation critical for both developing organs and adult vascular smooth muscle and endothelium; and (5) muscle-system processes. Collectively these terms describe a molecular program that is characteristically reactivated or dysregulated in adult vascular disease—including endothelial dysfunction, vascular smooth-muscle-cell (VSMC) phenotypic switching, pathological angiogenesis, and atherosclerotic remodeling.

### Biological Coherence of the Enrichment Signature

The top KEGG pathways map directly onto this developmental-reactivation theme. Cadherin signaling (especially VE-cadherin, N-cadherin, and protocadherin) is essential for endothelial barrier integrity, cell–cell adhesion during vasculogenesis, and pathological vascular remodeling. Calcium and MAPK signaling function as major downstream effectors of growth-factor receptors (VEGF, FGF, PDGF) that drive embryonic vascular development and are re-engaged in adult PAD and atherosclerosis. The “cytoskeleton in muscle cells” pathway points squarely at VSMCs, whose phenotypic plasticity is a hallmark of vessel-wall remodeling in PAD. Thus, the overall enrichment profile (cadherin + calcium + MAPK + cytoskeleton) constitutes a classic signature of developmental programs being co-opted by adult vascular pathology.

### Comparison with Prior EWAS and Tissue Studies

Our findings are directionally consistent with the large 2026 multi-cohort EWAS of blood methylation across carotid, coronary, and peripheral atherosclerosis (Ingold, ten Cate et al., JACC). That study identified thousands of CpG sites associated with peripheral atherosclerosis, many of which overlapped smoking and cardiometabolic signatures; the strongest loci included AHRR, F2RL3, PRSS23, and intergenic regions near ALPP/ALPG. In the present PAD-enriched cohort the same directional changes were observed, albeit with lower prominence, which is expected given that PAD constituted only ∼20% of the multi-vascular-bed cohort while our study was composed entirely of PAD patients. The plaque-focused work of Aavik et al. (2015) demonstrated genome-wide hypomethylation and reactivation of the 14q32 miRNA cluster—again aligning with the hypomethylation predominance we report. Macrophage promoter-methylation shifts observed in hyperlipidemic/diabetic hind-limb ischemia models (Babu et al., 2015) further support the concept that epigenetic reprogramming of inflammatory and angiogenic gene sets contributes to impaired vascular recovery in PAD.

Race and ancestry are important modifiers of the methylome. Stolze et al. (2025) showed that self-reported race is a major driver of methylation variation in PAD, with distinct differentially methylated CpGs within Black versus White strata. Because our pilot cohort was exclusively European ancestry, the present results should not be extrapolated to other populations without further study.

### Potential Biomarker and Therapeutic Implications

If validated, the developmental and cell-motility methylation signature could serve as a blood-based biomarker for PAD risk stratification or disease activity beyond traditional clinical factors. In addition, the observation that many of the same pathways are modulated by demethylating agents (e.g., 5-azacytidine) or lifestyle interventions that alter one-carbon metabolism raises the possibility of epigenetic therapies aimed at restoring a more physiologic methylation state and improving perfusion or limiting progression. Such approaches remain experimental and will require rigorous safety and efficacy testing in PAD-specific models and trials.

### Limitations

Several limitations must be acknowledged. First, the sample size is small (n = 10 PAD vs. 22 controls), rendering the study hypothesis-generating rather than confirmatory; the large number of nominally significant loci partly reflects this low statistical power after multiple-testing correction. Second, blood is a heterogeneous tissue; cell-type composition differences between PAD patients and controls could contribute to observed methylation differences, although SeSAMe and related pipelines mitigate some technical artifacts. Third, the design is cross-sectional, so causality cannot be inferred. Fourth, clinical covariates (smoking pack-years, diabetes severity, medication use, ankle-brachial index) were not fully captured for multivariable adjustment. Finally, the cohort was restricted to individuals of European ancestry, limiting generalizability.

## Conclusions

Genome-wide DNA methylation profiling of peripheral blood identifies a high-confidence set of loci and regions that are differentially methylated in PAD and strongly enriched for embryonic vascular and muscle developmental programs, cell adhesion/motility machinery, and calcium/MAPK signaling. These findings support the concept that epigenetic reprogramming of developmental gene networks contributes to the pathogenesis of PAD and provides a foundation for larger multi-ethnic validation studies, biomarker development, and exploration of epigenetic therapeutic strategies.

## Data Availability

All data produced in the present study are available upon reasonable request to the authors

## Acknowledgments

The authors thank the patients who participated in this study and the staff of the Penn State Hershey Vascular Surgery Clinic for assistance with recruitment. Methylation profiling was performed by TruDiagnostic Inc.

## Data Availability

De-identified methylation data (IDAT files and processed β-value matrices) will be deposited in a public repository (e.g., GEO) upon publication. Code used for SeSAMe processing and enrichment analysis is available from the corresponding author upon reasonable request.

## Declarations

Author Contribution Statement. PKJ, AS and VRV contributed to the conception/design of the study and contributed to data acquisition. AS, KS and CJ performed patient recruitment, consent, and sample acquisition, PKJ performed data analysis. PKJ, VRV and AS performed data interpretation, drafted the manuscript, and critically revised the manuscript for important intellectual content. All authors approved the final version of the manuscript and agree to be accountable for all aspects of the work.

## Ethics Approval and Consent to Participate

This study was conducted in accordance with the principles of the Declaration of Helsinki and was approved by the Penn State College of Medicine Institutional Review Board/Human Subjects Protection Office (IRB #21252EP). For PAD patients, eligibility screening was performed using electronic medical records and confirmed by clinician review. Given the acute nature of critical illness, informed consent was obtained from patients or their legally authorized representatives, as appropriate before enrollment. Samples from healthy control participants were obtained through a commercial genomic DNA assay conducted by Polbiomed LLC. All control participants provided explicit written informed consent for the collection of biological specimens and the use of their de-identified genomic data for research purposes. All data and biospecimens were de-identified prior to analysis to ensure participant confidentiality.

